# Gender dysphoria and sexual euphoria – A Bayesian perspective on the influence of gender-affirming hormone therapy on sexual arousal

**DOI:** 10.1101/2021.11.22.21266679

**Authors:** Manfred Klöbl, Murray Bruce Reed, Patricia Handschuh, Ulrike Kaufmann, Melisande Elisabeth Konadu, Vera Ritter, Benjamin Spurny-Dworak, Georg S. Kranz, Rupert Lanzenberger, Marie Spies

## Abstract

While the concept of sexual orientation is more clearly defined in cisgender, this is less so in transgender individuals. Both experienced gender and sex hormones have a relation to sexual preferences, arousal in response to erotic stimuli, and thus sexual orientation. In transgender individuals sexual orientation occasionally changes before or during transition, which may involve gender-affirming hormone therapy. Using functional magnetic resonance imaging, we investigated whether the neuronal and behavioral patterns of sexual arousal in transgender individuals moved from the given (before) to their chosen gender after 4.5 months of hormone therapy. To this aim, trans women and men as well as age-matched cisgender controls rated visual stimuli showing heterosexual, lesbian or gay intercourse for subjective sexual arousal. Utilizing a Bayesian framework allowed us to incorporate behavioral findings in cisgender individuals of different sexual orientations. The hypothesized changes in response patterns could indeed be observed in the behavioral responses to the single but not the differentiation between stimulus categories with the strongest results for trans men and lesbian scenes. Activation of the ventral striatum supported our hypothesis only for lesbian scenes in trans women. This prominent role of lesbian stimuli might be explained by their differential responses in cis women and men. We show that correlates of sexual arousal in transgender individuals might change in direction of the chosen gender. Future investigations longer into transition might resolve the discrepancy on behavioral and neuronal levels.

## Introduction

“Sex”, in the equivocation of the term, connects intercourse and chromosomally determined gender, which is most often congruent with given gender identity. This duality comes along with interrelations manifesting as sexual orientations as well as biological and social self-perceptions. The categorical concepts of hetero-, bi- and homosexuality are relatively clear for cisgender (CX) but much less so when considering transgender (TX) persons (Auer et al., 2014; Cerwenka et al., 2014; Laube et al., 2020; Schrock & Reid, 2006). Although the terms “androphilia”, “gynephilia” and “ambiphilia” are less ambiguous, they do not reflect the fluidity of sexual orientation. Moreover, sexual orientation in TX individuals may change before or during transition (Auer et al., 2014).

Correlates of sexual arousal have been shown to vary based on biological sex, which may correspond with given gender. CX men (CM) and women (CW) show differential responses to erotic stimuli, where CW exhibit lower levels of subjectively rated sexual arousal (Murnen & Stockton, 1997). Furthermore, CW show weaker relationships between sexual orientation and arousal as assed via genital and subjective measures as well as pupil dilation when viewing erotic stimuli (Rieger et al., 2015). Interestingly, homosexual CW presented an exception to this finding. An earlier investigation reported that the stimuli-specific genital and subjective response patterns differentiating preferred and non-preferred sex are similar for CM and post-operative trans women (TW, i.e., male-to-female TX individuals) (Chivers et al., 2004), supporting a role of biological sex in mediating arousal despite transition.

However, these differences between CM and CW are not necessary reflected on a neuronal level, as a meta-analysis of functional magnetic resonance imaging (fMRI) data has shown (Mitricheva et al., 2019). This apparent contradiction might be related to a desynchrony in genital and subjective arousal much stronger in CW than in CM (Meston & Stanton, 2019; Sierra et al., 2019). Furthermore, bisexual CM also show substantially more inter-individual variance in genital arousal patterns than hetero- or homosexual male subjects (Slettevold et al., 2019). Thus, comparative investigations of sexual arousal should consider subjective as well as objective measures to capture the full spectrum of responses.

Besides genital measures, for which an ascertainment bias needs to be taken into account (Chivers et al., 2004), neuronal responses might provide an objective way to quantify sexual arousal (Ponseti et al., 2006). However, to date insufficient emphasis has been placed on the confounding influence of sex hormones on brain function (for reviews see Heany et al. (2016); Slettevold et al. (2019)). Studying patients suffering from gender dysphoria receiving high doses of sex hormones as part of their gender-affirming hormone therapy (GHT) further offers the possibility to jointly investigate the role of sex hormones, biological and social gender on sexual arousal (Kranz et al., 2020). This model has been utilized to demonstrate an influence of sex, gender, gender dysphoria and / or sex hormones on various structural features of the brain (e.g., Baldinger-Melich et al. (2020); Flint et al. (2020); Hahn et al. (2015); Kranz et al. (2018); Zubiaurre-Elorza et al. (2014); Zubiaurre-Elorza et al. (2013); see Guillamon et al. (2016) for a detailed review). Many sex-related structural effects can be explained by overall brain size (Jäncke et al., 2015; Lotze et al., 2019), whereas social behavior was associated with regional gray matter volume (Kiesow et al., 2020) and task activation with strategic differences (Gaillard et al., 2021). Although none of these results support the picture of a clear dimorphism, they still highlight relationships between sex differences on behavioral and neuronal levels.

As for the neuronal correlates of sexual arousal, only the hypothalamus and the ventral striatum (VS) were found to be explicitly activated by erotic stimuli compared to general arousal (Walter et al., 2008). The hypothalamus has previously been related specifically to male sexuality (Brunetti et al., 2008; Karama et al., 2002). Based on these discoveries, it was shown that sex and sexual orientation not only influence the magnitude of the behavioral and VS response but also its specificity for the stimulation material. This partly explained ambiguities for bisexual subjects (Safron et al., 2020; Safron et al., 2017). However, since only CX individuals were included in these previous works, a differential consideration of sex and gender as driving factors was not possible. Moreover, longitudinal studies with TX people investigating the influence of GHT on the neuronal correlates of sexual arousal are generally missing. Unifying these open questions and building upon previous findings, we thus investigated whether behavioral and fMRI response patterns to erotic stimuli move from that of the given to the chosen gender over the course of GHT.

## Methods

This study was conducted according to the Declaration of Helsinki including all current revisions and the good scientific practice guidelines of the Medical University of Vienna. The protocol was approved by the institutional review board (EK Nr.: 1104/2016) and registered at clinicaltrials.gov (NCT02715232).

### Study design

The study followed a controlled longitudinal observational design. All participants underwent two MRI sessions with an interval of (median ± interquartile range) 138 ± 35 days. Following the first MRI, hormone treatment following an individualized regimen according to protocols of the Department of Obstetrics and Gynecology at the Medical University of Vienna was initiated. MRI sessions included structural, diffusion-weighted, task, resting-state and spectroscopy scans. Only the task data is presented here. Blood for determining testosterone, progesterone and estradiol plasma levels was drawn at the day of the MRI assessments. The Klein Sexual Orientation Grid (KSOG) was administered 14 ± 24 days before the first and 14 ± 23 days after the second MRI appointment. Missing hormone and KSOG values were estimated using the Missing Data Imputation (MDI) toolbox, version 2, in MATLAB (Folch-Fortuny et al., 2016) (see supplement for details).

### Participants

TX individuals seeking GHT were recruited from the Unit for Gender Identity Disorder, Department of Gynecology and Obstetrics at the General Hospital in Vienna. CX control subjects were recruited via social media, designated message boards at the Medical University of Vienna and from a Neuroimaging Labs database. CM and CW were age-matched ±3 years to TW and TM, respectively. Inclusion criteria comprised: a diagnosis of gender dysphoria (TX individuals only) according to the Diagnostic and Statistical Manual for Mental Disorders, version 5 (DSM-5: 302.85) or the International Classification of Diseases, version 10 (ICD-10: F64.1); general health based on medical history, physical examination, electrocardiogram, laboratory screening and structural clinical interview (SCID) for DSM-IV Axis-I. Participants were excluded in case of major neurological or internal illnesses, pregnancy, severe Axis-I comorbidities or any Axis-I disorder in CX, steroid hormone treatment within six months prior to inclusion, treatment with psychotropic agents three months prior to inclusion, clinically relevant abnormal laboratory values, MRI contraindications, current substance abuse (excluding nicotine), current or past substance-related disorder and insufficient compliance. All participants gave written informed consent.

### Sexual arousal task

In the MRI scanner, participants were visually presented a sequence of blocks with five explicit images of heterosexual 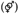, lesbian 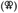 or gay 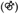 intercourse. Sports scenes with two women (SW) or two men (SM) constituted the control for general arousal. Each image was shown for 4 s. Participants were instructed to indicate whether they are “strongly turned on”, “turned on”, “turned off”, or “strongly turned off” by the erotic stimuli using fingers of their right hand and an MRI-compatible four-button response device. For the sports scenes, participants were instructed to indicate how much they liked the image since rating them for arousal led to confusion in pilot runs. Four smiley faces with strong positive to strong negative expressions were depicted below the stimuli and circled in red after pressing the respective button. The 20-s-blocks were flanked with baseline periods of the same length showing a fixation cross in the center of the stimulus frame. The task contained four blocks of each sexual arousal and two of each control condition in random order. Before entering the scanner, participants received written instructions with sample images and performed a short training run to familiarize with the experiment and degree of explicitness. In order to further encourage genuine answers, we assured participants that data will be anonymized before analysis and the experimenter is not watching the stimulation screen (the lid of the stimulation laptop was closed). The stimuli from Safron et al. (2007); Safron et al. (2017); Sylva et al. (2013) and normative scores were kindly provided by the authors. Since the material did not contain heterosexual scenes, we added these from a database of pornographic movie stills. This was done in order to prevent generalized aversive reactions to homosexual stimuli, which might be expected for the CW group (Sylva et al., 2013). An internal validation ensured comparable style and explicitness to the existing stimuli.

### FMRI acquisition, preprocessing and modeling

Scanning was conducted on a Siemens Prisma 3 T machine: TE / TR = 30 / 2050 ms, GRAPPA 2, 210 × 210 mm field of view, 100 × 100 pixel in-plane resolution, 35 axial slices of 2.8 mm (25% gap), flip angle 90°, orientation parallel to the anterior-posterior commissure line.

For data preprocessing, physiological artifacts were first reduced via PESTICA (Beall & Lowe, 2007). Subsequent steps were performed using SPM12 (https://www.fil.ion.ucl.ac.uk/spm/software/spm12/), unless otherwise specified. Slice-timing correction was performed to the temporally middle slice, followed by two-pass realignment of both measurements per subject to the mean image. Images were normalized to the standard space defined by the Montreal Neurological Institute (MNI) and resliced to 2.5 mm isotropic, approximately maintaining the voxel volume (K. Mueller et al., 2017). The BrainWavelet Toolbox (Patel et al., 2014) was used for nonlinear artifact reduction with the “chsearch” parameter set to “harsh” for increased artifact sensitivity and the “threshold” set to “20” due to the application to unsmoothed data with decreased signal-to-noise ratio by GRAPPA acceleration. Images were gray-matter masked using a custom template (Klöbl et al., 2020) and smoothed with a Gaussian kernel of three times the resliced voxel size.

Single subject models contained one regressor per condition 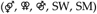, the Friston-24 model of motion (Friston et al., 1996) and an automatically derived number of combined white matter and cerebrospinal fluid regressors for an adapted CompCor approach (Behzadi et al., 2007; Klöbl et al., 2020). Contrasts for *raw* erotic 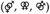 and sports scenes (SW, SM) as well as *category-specific* activations 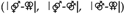 were calculated. Median activation was extracted from the VS region in the Automated Anatomical Labeling atlas 3 (AAL3) (Rolls et al., 2020) using the MarsBaR toolbox, version 0.44 (http://marsbar.sourceforge.net/). Despite its activation specific to sexual arousal, the hypothalamus was not investigated as target region due to its heterogeneous structure.

### Statistical modeling and inference

Bayesian multivariate multilevel models and nonlinear hypothesis testing (brms_2.15.0 R package (Bürkner, 2017) run in RStudio 1.4.1717) were chosen as statistical framework for our analyses for the following reasons: First, we aimed to incorporate previous behavioral results from comparable studies. Second, the hypotheses including absolute differences are not easily evaluable in many other approaches. Third, the posterior probabilities provided by Bayesian statistics facilitate more fine-grained interpretations, matching also the spectrum perspective on gender. Bayesian multilevel models further protect against multiplicity issues (Gelman et al., 2012) allowing for more in-depth inferences. It was first tested whether the responses of all TX participants over all sexual arousal conditions moved from the given to the chosen gender:

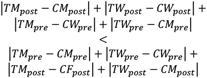

However, neither similar behavior between TW and TM nor across conditions can be assumed. Thus, sub-hypotheses separating the TW and TM as well as the stimuli were further evaluated.

We first investigated VS and behavioral *category specificities* between the conditions since the absolute differences in elucidated responses were shown to differ between sexes and orientations (Safron et al., 2020; Safron et al., 2017). Subsequently, since no particularly strong evidence was found for changes in *category specificity*, the *raw* responses of all five conditions were considered to unveil whether the absence of more specific effects resulted from generally comparable responses, added variance or a general lack of effects.

The sexual preference of the participants at each time point was quantified as the mean of the KSOG items “current sexual attraction”, “current sexual behavior” and “current sexual phantasies” which were rated on a seven-point Likert scale from “women only” to “men only”. The score was mean-centered per group and time point and used as covariate in all models to account for the effects on orientation-specific sexual arousal. The same was done for participant age to avoid collinearity due to differences between the groups. To correct for the varying GHT regimens, the first principal components of the standardized log-transformed testosterone, progesterone and estradiol levels were included as separate covariates for TM and TW at the second measurement since group-specific effects were likely (S. C. Mueller et al., 2020). The models further contained fixed group and time point factors, their interactions and a random intercept per participant with correlations between the conditions. Priors for behavioral data were calculated based on the sexual orientation score, the results of Gizewski et al. (2009); Ku et al. (2013); Safron et al. (2007) and the normative data from Safron et al. (2007) kindly provided by the authors (see supplement for mathematical details). Pairwise comparisons of the factors give 95% highest posterior density (HPD) regions, i.e., areas of the posterior distributions containing 95% of the estimated samples. The nonlinear hypothesis tests return posterior probabilities (PP) of support by the data.

## Results

The demographics and hormone levels of the study sample are provided in Table 1. Less TW (and matched CM) could be recruited compared to TM and CW. Due to age matching, the former groups are also older. Heterosexual stimuli were received as turning on over all groups, whereas gay stimuli were received as turning off. The lesbian stimuli show a marked difference between CM and CW. At the second assessment, TW had numerically lower testosterone and higher progesterone and estradiol levels, whereas TM had higher testosterone, lower progesterone and lower estradiol levels. The hormone levels of the CX groups were more stable with slight variations in progesterone as well as estradiol in CW only.

**Table 1:**
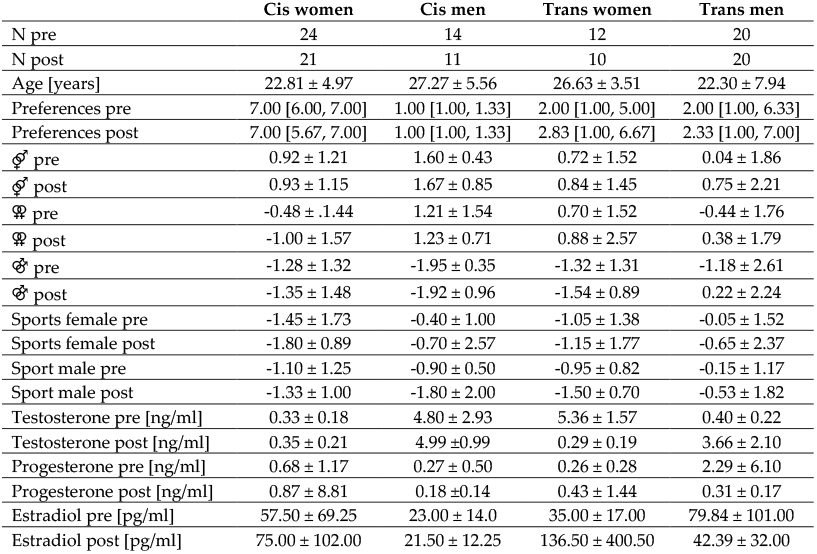
Demographics and covariates. Median values and interquartile ranges (third minus first) are given, except for the sexual preferences where minimum and maximum are provided for being more informative. Age is given at the first measurement. Sexual preferences were calculated from the Klein Sexual Orientation Grid with 1 = women only and 7 = men only and is provided as median [minimum, maximum]. The interval of the behavioral responses to the sexual stimuli was −2 = strongly turning off to 2 = strongly turning on. The scores are not corrected for sexual orientation. Hormone levels for the cisgender participants and pre-treatment time points are provided for comparison only and were not included in the statistical analysis.

The behavioral model on *category-specific* data shows a generally stronger differentiation to gay stimuli for the CM (visualized for 95%-HPDs not covering zero in Figure 1). The differentiation between heterosexual and gay stimuli decreased for the TM. However, the most differentiation between lesbian and heterosexual stimuli was found for the CW and it was also stronger in TW than TM. Weak to moderate support for our hypothesis of TX response patterns to sexual arousal moving from the given to the chosen gender one was found for the *category-specific* behavioral data (PPs not exceeding 80%). This was still strongest in TW for the sub-hypothesis on differences between lesbian and gay and overall weak for the difference between heterosexual and gay stimuli. However, the *raw* behavioral data clearly showed the hypothesized changes. This effect was again especially strong for lesbian stimuli but also in TM as indicated by the highest PPs. An overview of the behavioral results is given in Figure 1. Moderate to strong correlations of random intercepts were found for raw heterosexual and lesbian (r = 0.57, 95% credible interval [0.32, 0.76]), heterosexual and female sports (r = 0.36, [0.00, 0.68]), lesbian and female sports (r = 0.41, [0.05, 0.72]) and female and male sports scenes (r = 0.73, [0.33, 0.95]; see supplement for full data).

**Figure 1:**
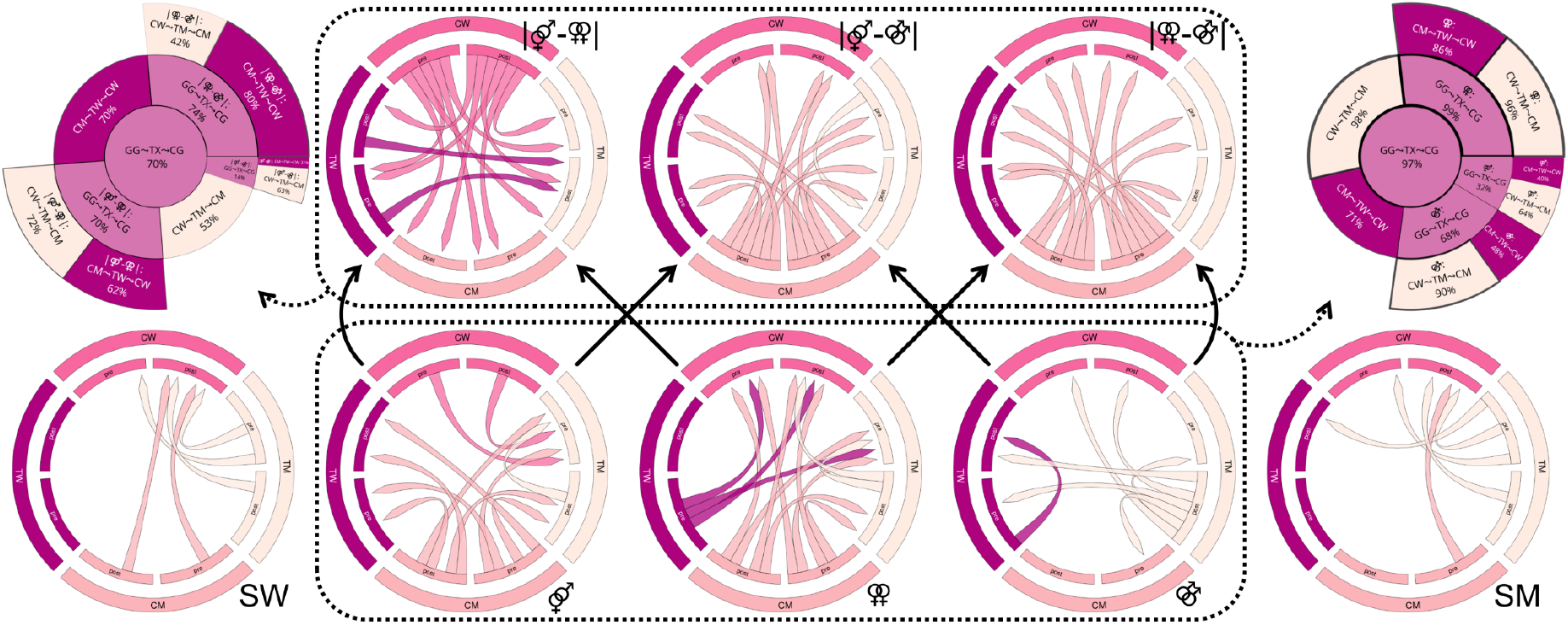
Results of the behavioral analyses. For visualization purposes, the chord diagrams only show the differences between groups and time points with a 95% highest posterior density not covering 0. The top row shows the condition-specific, the bottom row the raw differences. Arrow directions indicate “greater than” relations. The hypothesis tests for response patterns distancing from the given and approaching the chosen gender are presented on the top left (condition-specific responses) and right (raw responses) with their respective posterior probabilities. The outer sectors represent sub-hypotheses of the inner sectors and central main hypothesis. The results for the sports scenes showing women (SW) or men (SM) are presented for comparison and were not included in the hypothesis tests. GG: given gender, TX: transgender, CG: chosen gender, TW: trans women, TM: trans men, CW: cis women, CM: cis men, pre: pre-treatment assessment, post: post-treatment assessment.

The *category-specific* effects were generally weaker for the VS activation, which was only clearly smaller for the TM after 4.5 months of GHT compared to the other groups (see 95%-HPDs in Figure 2). Distinct changes in the *raw* data according to our hypotheses were found mostly for TW (PP = 90%), especially with lesbian stimuli (PP = 97%). An overview of the results for the ventral striatum activation is given in Figure 2. Correlations between the categories were weaker with considerably larger credible intervals. Complete details on the models and tests are provided in the supplementary tables.

**Figure 2:**
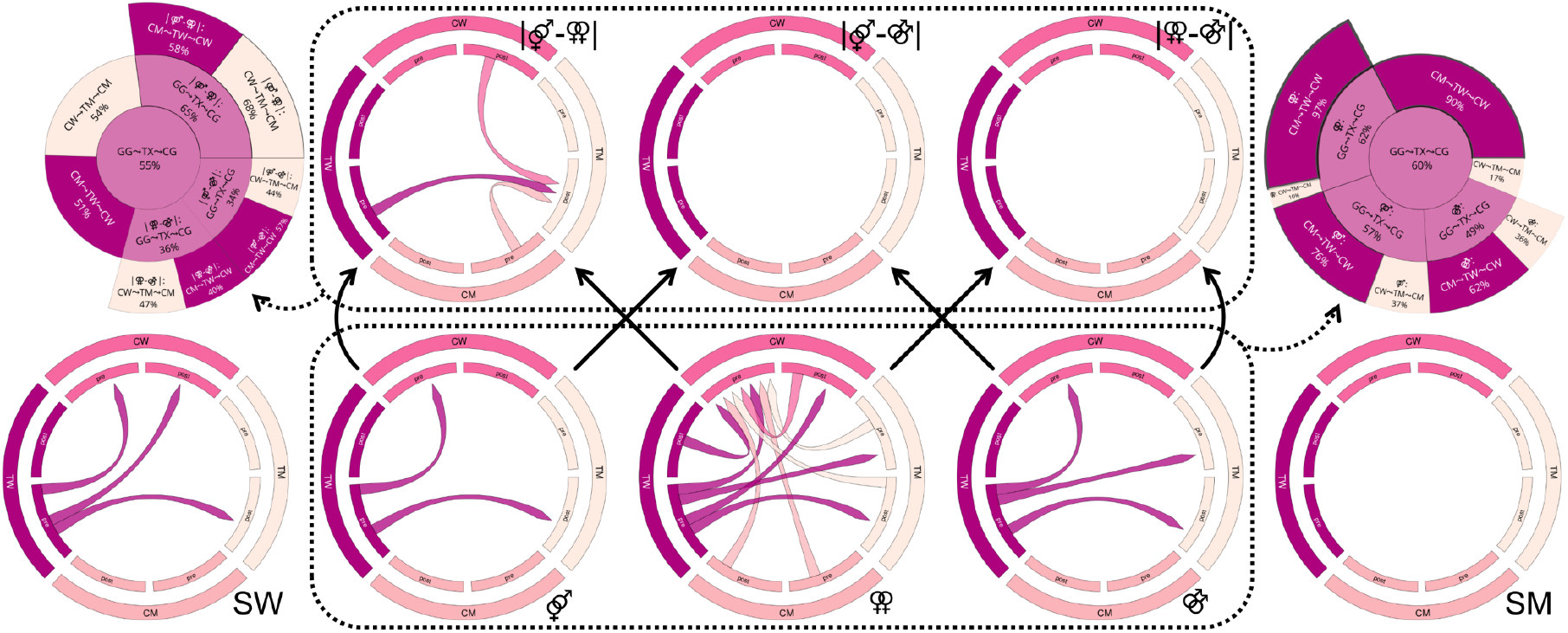
Results of the ventral striatum activation analyses. For visualization purposes, the chord diagrams only show the differences between groups and time points with a 95% highest posterior density not covering 0. The top row shows the *condition-specific*, the bottom row the *raw* differences. Arrow directions indicate “greater than” relations. The hypothesis tests for activation patterns distancing from the given and approaching the chosen gender are presented on the top left (*condition-specific* activation) and right (*raw* activation) with their respective posterior probabilities. The outer sectors represent sub-hypotheses of the inner sectors and central main hypothesis. The results for the sports scenes showing women (SW) or men (SM) are presented for comparison and were not included in the hypothesis tests. GG: given gender, TX: transgender, CG: chosen gender, TW: trans women, TM: trans men, CW: cis women, CM: cis men, pre: pre-treatment assessment, post: post-treatment assessment.

## Discussion

We investigated whether the behavioral and neuronal responses to sexual arousal in TX individuals move from the given to the chosen gender over the course of 4.5 months of GHT. Overall strong support for this hypothesis was found for the *raw* behavioral responses, especially for the TM group and lesbian scenes. For VS activation a comparable effect was observed for these stimuli presented to the TW group. The support for *category-specific* changes was markedly weaker as indicated by the lower PPs.

The cissexual groups showed markedly stronger *category-specific* differentiation in behavioral responses. Extending previous work we also included heterosexual stimuli to avoid receiving only aversive responses from CW (Sylva et al., 2013). Indeed, the most behavioral differentiation was found for heterosexual vs. lesbian scenes in CW and heterosexual or lesbian vs. gay scenes in CM. These findings add to the results from Safron et al. (2020); Safron et al. (2017) insofar as that they also show strong differentiation for the CW group, which, on average, signaled subjective arousal only for the heterosexual stimuli. Overall, the behavioral response to the heterosexual stimuli was still the most similar between CW and CM subjects.

### Effects in trans men

A remarkable change in *category specificity* as well as spanning all *raw* responses to erotic stimuli was observed for the TM group: As the subjective arousal for all stimuli categories increased, with a prominently higher reported sexual arousal for gay scenes than all other groups, the specificity for especially heterosexual vs. gay stimuli decreased. Furthermore, the VS activation showed less *category specificity* after GHT compared to the other groups. The *raw* increases were absent in the neuronal data. Altogether, these results provided at most weak support for our hypothesis on *category-specific* responses in TM but might point towards generally higher subjective sexual arousal. Even among androphilic and ambiphilic / bisexual transitioned TM, there might be a stronger expression of bisexualism than in CM of the same reported orientations (Bockting et al., 2009). In a mixed pre- and post-surgical TM sample, measures of genital arousal displayed traits of CW and CM controls (Raines et al., 2021): Although the male-typical preference of orientation-congruent stimuli was observed, there was still the more female-typical response to all stimuli. Our sample identified as mostly gynephilic, but preset bisexual rather than monosexual traits could partly explain the lack of clear changes in *category-specific* responses to sexual arousal. Of note, the TM also had the least aversive behavioral responses to the sports scenes. Together with the correlations found between erotic and sports stimuli, this might raise questions on the perception of general arousal. However, as no strong pre-post changes for the sports scenes were identified, as well as no changes towards the response pattern of the chosen gender (see supplementary tables), sufficient specificity of the responses to the erotic stimuli is assumed.

Indeed, *raw* behavioral responses provide general support for our hypothesis, particularly with the TM pattern becoming more CM- / less CW-typical across erotic stimuli of all orientations. In detail, the increase in arousal for lesbian scenes can be explained by the response approaching a CM and for the gay by distancing from a CF pattern. On the contrary, the VS activation does not show any changes in the hypothesized direction. This stark difference between behavioral and neuronal measures offers two explanations: First, the activation for sexual arousal does not change in the VS for TM after GHT. Second, there is incongruence between subjectively perceived and neuronally controlled level of arousal. In an fMRI study with accompanying behavioral data, a small sample of gynephilic-leaning post-operative TM, as expected, showed a significantly higher level of reported arousal to pictures showing two nude women compared to nude men (G.-W. Kim et al., 2016; T. H. Kim et al., 2016). Activation of the hypothalamus, but not the VS was reported. Where the hypothalamus was found, like the VS, to react to sexual rather than general arousal (Walter et al., 2008), this reaction is potentially sex-specific and predominantly found in men (Brunetti et al., 2008; Karama et al., 2002). While the hypothalamus was not investigated here, this might be done in future studies on a voxelwise basis or after refining the target region (Osada et al., 2017), keeping structural sex differences in mind (Makris et al., 2013).

### Effects in trans women

The TW group showed stronger behavioral *category specificity* for heterosexual vs. lesbian scenes compared to the TM group, which resembles the CW. This effect is also found for the VS activation but only for pre-treatment TW and post-treatment TM. Pre-treatment, the TW’s *raw* response to lesbian stimuli was more positive than that of the CW or the TM group before therapy, establishing a certain similarity to the CM group. This adds to the generally strong support of our hypothesis for lesbian stimuli also observable for the TW. Contrary to the TM, the TW showed increased VS activation for all conditions except the sports scenes depicting men, clearly standing out from the other groups even more than the CM. This general level of VS activation also explains the lack of differentiation in neuronal responses. Despite weaker adaptations on the behavioral level, the neuronal data evidently reveals changes from the given towards the chosen gender in TW, strongest for sexual arousal through lesbian scenes.

In a study on post-operative TW, premenopausal and menopausal CW, similar activation patterns to nude pictures were found in the former two but not in the last group (Kim & Jeong, 2014). These results need to be interpreted with caution due to the age difference between groups (despite correction) and its highly exploratory nature but point towards similarities in TW after transitioning and CW, as well as hormonal influences on sexual arousal. A large fMRI study comparing TX individuals living in their chosen gender to CX controls reported significantly lower activation for erotic stimuli in the right superior temporal gyrus for TW as compared to CM (S. C. Mueller et al., 2020). The investigation was based on the “neurophenomenological model of sexual arousal” proposed by Stoléru et al. (2012). According to this model, the superior temporal gyrus belongs to the “inhibition and devaluation” component, the VS to the “motivational” component (as the hypothalamus). Interestingly, for all groups, the highest interhemispheric task connectivity for the superior temporal gyrus was also found in TW. Given the relationship between VS activation and whole-brain functional connectivity (Mori et al., 2019), seed-based analyses might shed further light on its particular importance for sexual arousal in TW.

The communalities in behavioral and neuronal responses (i.e., both being strongest for lesbian stimuli) might be related to the controversial concept of autogynephilia in TW (Blanchard, 1989). While the self-reported orientation of our TW sample is gynephilic on average before and after GHT (Auer et al., 2014), the perceived sexual arousal by watching the intercourse of other women might have experienced a potentially hormone-induced actualization. A further point that needs consideration is the more negative experience of sexuality in TW (Gil-Llario et al., 2021). While this might not introduce a particular bias unless TW are directly compared to TM, influences on the neuronal and behavioral correlates of sexual arousal cannot be ruled out and might have contributed to the lack of behavioral effects.

## Limitations

Less TW could be recruited and a considerable amount of TW dropped out of the study, which is in line with the reported incidence of gender-confirming surgery (Nolan et al., 2019). Even though this seems to be a general trend, there are societal and age-related influences on the ratio of gender dysphoria in biological males and females. The higher estimated prevalence of gender dysphoria in biological males might be driven by the more complex developmental sex differentiation processes on a hormonal but also psychosocial level (Zucker, 2017). However, the psychosocial factors might also lead to a distortion of the true prevalence ratio. In light of our results, this might however not have led to an underpowered sample. Still, simplifications were necessary when correcting for the potential influence of the varying treatment regimens. Since this study on TX individuals rather than sexual preferences, mixed orientations are found especially in the TW and TM groups. Also, some CX participants might be described as heterosexual-leaning rather than strictly heterosexual. Correcting for the influence of varying sexual orientation leads to results for average responses which are gynephilic-leaning in both TX groups. Lastly, it is not possible to conclude whether the effects found solely stem from the influence of the hormones on brain and behavior or result from an increased self-perception of the chosen gender or potential social influences during transition. Also we assumed categorical, not fluid gender, i.e. the extent to which someone may experience themselves as belonging to a certain gender would affect experienced sexual orientation.

## Conclusion

In line with our hypothesis, we propose that *raw* behavioral and neuronal measures of sexual arousal in TX individuals might approach the pattern of the chosen gender during GHT. These effects were considerably stronger for erotic stimuli of a certain orientation than the *category specificity*. The underlying reasons might be that the responses to different stimulus categories changed in the same direction or that the diverse sexual orientations of the participants as well as the contrasting of the responses introduced additional variance. Where behavioral analyses showed a generally strong support for our hypothesis (slightly stronger in TM), this was only the case for the VS activation only in TW. The most prominent changes were found for lesbian scenes. While we cannot offer a definite explanation for this observation, the general difference between CM and CW responses could provide increased sensitivity to changes for this category. In TW, the controversial concept of autogynephilia could also offer a perspective. Future studies in TX individuals could extend their focus to the hypothalamus, which constitutes a promising target given its role in male sexuality. Furthermore, in-depth analyses of the role of the sexual orientation in the transitioning process would shed more light on changes in perceived sexuality.

## Supporting information

Supplementary methods and results

Results for differential responses

Results for raw responses

Results for differential ventral striatum activation

Results for raw ventral striatum activation

## Data Availability

Due to reasons of data protection, the preprocessed data is available only upon reasonable request to the corresponding author.

## Code availability

Software for data processing and analysis was used as indicated. The code is available from the corresponding author upon reasonable request.

## Author contributions

The study was planned by Rupert Lanzenberger and Georg Kranz. The fMRI task presented here was conventionalized by Georg Kranz and Manfred Klöbl. Magnetic resonance imaging data was acquired by Manfred Klöbl, Murray Bruce Reed and Benjamin Spurny-Dworak. Clinical data was acquired by Marie Spies, Patricia Handschuh and Melisande Elisabeth Konadu. Ulrike Kaufmann, Vera Ritter and Melisande Elisabeth Konadu managed subject recruitment and handling. Ulrike Kaufmann further provided medical support. Marie Spies and Patricia Handschuh supervised the study. Manfred Klöbl planned and conducted the statistical analysis and wrote the first draft of the manuscript. Georg Kranz and Marie Spies provided methodological supervision. All authors have contributed to the interpretation of the data, critically read and revised the manuscript.

## Acknowledgements

We thank the students of the Neuroimaging Labs for their continuous support throughout the study. We are grateful towards the Center of Excellence for MR Research at the Medical University of Vienna for the received support and supply with consumables.

