## Supplementary methods and results for "Gender dysphoria and sexual euphoria – A Bayesian perspective on the influence of gender-affirming hormone therapy on sexual arousal"

### Sex hormones

Luteinizing hormone, follicle-stimulating hormone, progesterone, estradiol, testosterone, sex hormone binding globulin and dehydroepiandrosterone sulfate were determined from blood drawn at the days of both MRI session. While only testosterone, progesterone, and estradiol were used in the analyses, the other hormones were used for imputation purposes.

### Missing data imputation

As recommended for the given data structure, trimmed-scores regression (TSR) with the standard settings of the MDI toolbox was used (Folch-Fortuny et al., 2015). The number of components with the minimum predicted residual sum of squares (PRESS) was retained. Since the hormone data was strongly right skewed and to avoid unreasonable negative estimates, imputation of missing values (CW: 8.33%, CM: 20%, TW: 23.50%, TM: 17.22%) was performed on log-transformed values. Back-transformed values were thresholded at 1.5 times the interquartile range from the first and third quartile of the original data to remove strong outliers. Due to distinct sex- and treatment-specific patterns with dependencies between first and second assessment, the time points were treated as different variables and imputation was performed for each group separately. Missing values in the KSOG (CW: 21.22%, CM: 20.18%, TW: 16.74%, TM: 14.01%) were treated similarly but rescaled and logit-transformed for imputation. To avoid infinite values, the interval [0, 1] was compressed to [1/(2N), 1-1/(2N)], where N is the number of elements in the complete dataset.

### Specifications of the Bayesian multilevel model

The outcome of the VS *response specificity* model was approximated using gamma distributions since the more appropriate folded normal or t-distributions were computationally not feasible and consistently produced divergent transitions in the Markov chains during distribution sampling. Normal distributions were used for the *content-specific* and *raw* activation models. The behavioral responses were originally coded {-2, -1, 1, 2} but rescaled to [0, 1] for statistical analysis. Since the absolute value of any difference on this scale is again on this scale, all behavioral data was modeled using zero-one-inflates beta distributions. Each model was run with 10000 iterations and 4 Markov chains. Convergence was checked visually and via the RHAT statistic. The "adapt_delta" parameter was set to "0.99" to eliminate divergent transitions.

### Prior calculation

Where possible, mean and standard deviation for the different subgroups and conditions were taken from Gizewski et al. (2009); Ku et al. (2013); Safron et al. (2007) (read from graphs if no exact values were found) as well as the normative data provided by the authors of Safron et al. (2007). The values are presented in Table S1 rescaled to the interval [0, 1], which

| Condition | Group | Orientation | Mean | Std | Reference/comment |
|---|---|---|---|---|---|
| ⚤ | CW | hetero | 0.78 | 0.09 | Gizewski, et al. (2009) |
| ⚤ | CW | bi | 0.73 | 0.12 | Ku, et al. (2013) (mixed sex) |
| ⚤ | CW | homo | 0.68 | 0.12 | extrapolated from hetero and bi |
| ⚤ | CM | hetero | 0.80 | 0.10 | Gizewski, et al. (2009) |
| ⚤ | CM | bi | 0.73 | 0.17 | Ku, et al. (2013) (mixed sex) |
| ⚤ | CM | homo | 0.66 | 0.17 | extrapolated from hetero and bi |
| ⚤ | TM | hetero | 0.72 | 0.17 | Ku, et al. (2013) (before treatment, due to similarity to Gizewski, et al. (2009) mixed group used for FtM, all orientations) |
| ⚤ | TM | bi | 0.72 | 0.17 | Ku, et al. (2013) (before treatment, due to similarity to Gizewski, et al. (2009) mixed group used for FtM, all orientations) |
| ⚤ | TM | homo | 0.72 | 0.17 | Ku, et al. (2013) (before treatment, due to similarity to Gizewski, et al. (2009) mixed group used for FtM, all orientations) |
| ⚤ | TW | hetero | 0.72 | 0.11 | Gizewski, et al. (2009) (before treatment; sexual orientation not differentiated in original publication, used for all) |
| ⚤ | TW | bi | 0.72 | 0.11 | Gizewski, et al. (2009) (before treatment; sexual orientation not differentiated in original publication, used for all) |
| ⚤ | TW | homo | 0.72 | 0.11 | Gizewski, et al. (2009) (before treatment; sexual orientation not differentiated in original publication, used for all) |
| ⚢ | CW | hetero | X | X | normative table |
| ⚢ | CW | bi | X | X | normative table |
| ⚢ | CW | homo | X | X | normative table |
| ⚢ | CM | hetero | 0.85 | 0.06 | Safron, et al. (2007), checked with normative table |
| ⚢ | CM | bi | X | X | normative table |
| ⚢ | CM | homo | 0.14 | 0.08 | Safron, et al. (2007), checked with normative table |
| ⚣ | CW | hetero | X | X | normative table |
| ⚣ | CW | bi | X | X | normative table |
| ⚣ | CW | homo | X | X | normative table |
| ⚣ | CM | hetero | 0.12 | 0.06 | Safron, et al. (2007), checked with normative table |
| ⚣ | CM | bi | X | X | normative table |
| ⚣ | CM | homo | 0.88 | 0.06 | Safron, et al. (2007), checked with normative table |
| SW | CM | hetero | 0.50 | 0.12 | Safron, et al. (2007) |
| SW | CM | bi | 0.36 | 0.12 | interpolated from hetreo and homo |
| SW | CM | homo | 0.21 | 0.12 | Safron, et al. (2007) |
| SM | CM | hetero | 0.50 | 0.08 | Safron, et al. (2007) |
| SM | CM | bi | 0.45 | 0.17 | interpolated from hetreo and homo |
| SM | CM | homo | 0.40 | 0.17 | Safron, et al. (2007) |

**Table S1: Mean and standard deviation of the responses for the different stimulus conditions per group.** Values that were only derivable from the normative data provided by Safron, et al. (2007) are not given since the data is not publicly available. The values were mapped from the original scales to [0, 1].

is the range of the beta distribution used for modeling the responses. Since the participants in Gizewski et al. (2009); Ku et al. (2013), other than in Safron et al. (2007), only had to indicate positive arousal, the lowest score was treated as 0.5 there. Based on the sexual preference score derived from the KSOG and the gender assigned at birth, the expected responses for each participant and condition in the current study were calculated. For preferences lying between homo- or hetero- and bisexuality, the responses were interpolated using the modified Akima approach in MATLAB (Akima, 1970). The average of all participants per group was used as prior mean. In order to avoid producing overly narrow priors, the maximum of the standard deviations for the single participants was used. If the sexual preference score fell between orientations, also the maximum of the two neighboring standard deviations was used. The priors for the difference scores were calculated as differences between the distributions (i.e., difference of means, sum of variances). Transformations from a normal to the logit scale of the beta distribution parameter priors were conducted according to Pedersen (2007).

**Supplementary results**

The complete model and test statistics are provided in the supplementary Excel tables. For interpretability in R, the following naming conventions were used: XH heterosexual stimuli, XL, lesbian stimuli, XG: gay stimuli, SW: sports stimuli showing women, SM: sports stimuli showing men, XHXG: absolute difference between heterosexual and gay stimuli, XHXL: absolute difference between heterosexual and lesbian stimuli, XLXG: absolute difference between lesbian and gay stimuli, G: group (i.e., {CW, CM, TW, TM}), M: measurement (i.e., {1 = pre-treatment, 2 = post-treatment}), Age0: age mean-centered per group, sex_present0: present sexual orientation score calculated from the KSOG and mean-centered per group, twPca: first principal component of standardized log-transformed post-treatment hormone levels for the TW group, tmPca: first principal component of standardized log-transformed post-treatment hormone levels for the TM group, GG: given gender, CG: chosen gender, TX: transgender.
